# Disruption of Anticipatory Insulin Secretion in Patients with Hyperglycemia and Obesity

**DOI:** 10.64898/2026.10.27.26366633

**Authors:** Doron Kleiman, Itia Magenheim-Samuel, Sarah Knapp-Goldin, Sienna Walsh, Renee Richens, Krisztina Toth, Jennifer Evans, Giang M Dao, Clinton R Bruce, Christopher S Shaw, Andrew C Betik, Vicky Kuriel, Shlomi Kotler, Greg M Kowalski, Tania Markovic, Malgorzata Brzozowska, Jerry R Greenfield, Rachel Ben-Haroush Schyr, Dorit Samocha-Bonet, Danny Ben-Zvi

## Abstract

Pancreatic beta cells integrate numerous signals in addition to glucose to control insulin secretion rate (ISR). Anticipatory insulin secretion, in which insulin is secreted according to future glucose levels through cephalic and hormonal cues can be effective in minimizing glycemic excursions. We show that in healthy individuals, ISR profile was best correlated with the future blood glucose profile (6±1 minutes) throughout the postprandial period, demonstrating anticipatory insulin secretion. The glucagon-like peptide-1 (GLP-1) profile was shifted by 9±3 minutes compared to the blood glucose profile, providing beta cells with information on future glucose levels via a feed forward mechanism. Obesity and pre-diabetes were associated with disruption of anticipatory insulin secretion: The GLP-1 and ISR profiles declined prematurely, contributing to prolonged post-prandial hyperglycemia. Agonism of the GLP-1 Receptor by semaglutide increased ISR and curbed hyperglycemia. However, semaglutide treatment abolished the anticipatory insulin secretion altogether: ISR and glucose profiles overlapped, resulting in a delay between insulin and glucose levels which was associated with higher post-prandial glycemic undershoots. Our data reveal that dysglycaemia disrupts the fine-tuned anticipatory insulin secretion, contributing to glycemic excursions in obesity, prediabetes and type 2 diabetes.

## Introduction

Pancreatic β cells secrete insulin in response to a rise in blood glucose levels, promoting glucose uptake and storage and maintaining glucose homeostasis. Insulin secretion rate (ISR) is modulated by incretin hormones (GLP-1 and glucose-dependent insulinotropic polypeptide, GIP), neural signaling, metabolites, intra-islet signals, intracellular regulators, and metabolic intermediates(1–6). The dynamics of ISR is further complicated by heterogeneity across individuals, islets, and β cells(7–11). Insulin clearance rate is regulated by insulin, and varies across tissues and physiological states(12–17). Consequently, the plasma insulin profile reflects the complex interplay between insulin secretion and clearance, both of which are affected by glucose and many other physiological factors.

The gut-secreted incretins, GLP-1 and GIP, are primary activators of the amplifying pathway in β cells, increasing ISR in response to glucose. Boosting ISR, alongside induction of weight loss underlies the activity of GLP-1 receptor agonists (GLP-1RAs) such as semaglutide, which improve glycemia in patients with type 2 diabetes(2, 18–21).

The core regulation over blood glucose levels relies on the negative feedback between glucose and insulin. Homeostatic feedback control systems have an inherent delay between the disturbance and the control signal(22), which can be overcome by feed forward control(23). In the insulin-glucose dynamics, a delay is introduced by insulin clearance rate, with insulin having a half-life of approximately 5 minutes(16). Feed-forward regulation would manifest as a temporal lead of ISR over glucose, i.e. anticipatory insulin secretion, and was suggested to be mediated in part by GLP-1.

Obesity and insulin resistance are associated with a disruption in β cell biology and impaired control of endogenous glucose production and peripheral tissue glucose uptake. β cells adapt to this challenge, leading to elevated fasting insulin, and an increase in β cell mass (9, 10, 24–28). Obesity and dysglycaemia affect the dynamics of ISR, and are associated with slower glucose uptake, blunted incretin effect, and lower insulin clearance(12–17).

Here, we quantify glucose, insulin, ISR and GLP-1 dynamics in healthy individuals and demonstrate anticipatory ISR throughout the postprandial period with GLP-1 acting as a feed-forward signal. As a result, the insulin and glucose profiles overlap. Obesity and dysglycaemia disrupt anticipatory insulin secretion, and are associated with a wider gap between the GLP-1 and glucose profiles, and a premature decline in ISR. Constitutive agonism of GLP-1 signaling by semaglutide improves glycemia and increases ISR, but may override endogenous feed-forward signals, thereby abolishing anticipatory ISR. In these patients, ISR and glucose levels overlap, causing a measurable delay between plasma insulin and glucose concentration profiles.

## Methods

The study was approved by the St Vincent’s Hospital Human Research Ethics Committee, Sydney, Australia (approval numbers 2019/ETH12216 and 2020/ETH02405). All participants provided written informed consent before the commencement of any study-related procedures.

Participants: Healthy participants were men and women aged 18-60 years who had a normal weight (BMI≤25 kg/m²), normo-glycemia, not treated with glucose-lowering medications without any underlying chronic condition. Participants in the obesity/dysglycaemia group were people living with obesity (BMI ≥ 30 kg/m²), with or without dysglycaemia, as well as people without obesity who had prediabetes or type 2 diabetes^29^, who had been advised by their managing physician to commence semaglutide treatment for an underlying condition. Initiating treatment with semaglutide was not mandatory, and not all participants commenced treatment with semaglutide during the study period. Exclusion criteria were pregnancy or lactation, current or previous smoking, alcohol consumption exceeding one standard drink per day, abnormal liver or kidney function, and treatment with medications known to affect glucose metabolism. See Supporting Information for further details.

Study visit: Following a screening visit to assess eligibility, participants attended the Clinical Research Facility following a 10-12-hour overnight fast to undergo a modified glucose challenge test. On arrival, weight, height, waist and hip circumference, and blood pressure were measured. An intravenous cannula was inserted into the antecubital vein, and the cannulated arm was kept warm using a heating blanket. Two baseline blood samples were collected 15 minutes apart. Participants then consumed a 50 g glucose solution (200 mL) within 5 minutes, followed by serial blood sampling for 3.5 hours (every 5 minutes during the first hour, every 10 minutes during the second hour, and every 15 minutes for the remainder of the study period). Participants who commenced treatment with semaglutide underwent an identical study protocol following treatment. Participants treated with glucose-lowering medication followed a washout protocol. We aimed to repeat the study 5-7 months from treatment commencement. See Supporting Information for a detailed protocol.

Biochemical Analysis: Plasma insulin, C-peptide and GLP-1 concentrations were measured using commercial ELISA kits, and glucose using the Nova StatStrip glucose monitoring system. C-peptide concentrations were used to calculate ISR using the ISEC deconvolution software program(29). See Supporting Information for further details.

Statistics: One-sample Student’s t-test was used to determine the difference from zero, and two-sample Student’s t-test to compare the means of two groups. Data are shown as mean and standard error of the mean (SEM).

Large Language Models were used for writing and formatting.

Data and Resource Availability: Deidentified participant data that underlie the findings reported in this article will be available immediately following publication, ending 5 years following the article publication, to researchers who provide a methodologically sound proposal to achieve the aims reported in the approved project proposal. Data may be obtained from the corresponding authors.

## Results

### Dynamics of ISR, insulin and glucose in healthy individuals

28 healthy individuals (Table 1) ingested 50 g of glucose following an overnight fast. Blood samples were collected every 5 minutes for the first hour, and at a lower frequency later for a total time of 210 minutes to capture glucose and hormones dynamics in high-resolution. We calculated ISR via deconvolution of the measured C-peptide profile(29) and compared it to the corresponding glucose profile (Figure 1A, Figure S1A). Both mean ISR and glucose levels rose within 5 minutes of the glucose ingestion; however, the mean maximal ISR occurred earlier (t=30 minutes) than the mean maximal glucose concentration (t=40 minutes).

**Table 1:** Characteristics of the healthy participants and participants with obesity/dysglycaemia.

| condition | units | Healthy |  | obesity/dysglycaemia |  | Healthy vs obesity/dysglycaemia | statistical test |
| --- | --- | --- | --- | --- | --- | --- | --- |
|  |  | Mean | Standard dev. | Mean | StDev |  |  |
| Age | years | 29.3 | 8.5 | 51 | 6.2 | **4.34E-15** | # |
| Gender | % male (n) | 46.4 (13) |  | 38.9 (14) |  | 7.26E-01 | & |
| Height | m | 1.7 | 0.1 | 1.7 | 0.1 | **1.82E-02** | # |
| Weight | kg | 64.3 | 7.6 | 95.6 | 21.9 | **3.57E-10** | # |
| BMI | kg/m^2 | 21.7 | 1.4 | 34.6 | 8.3 | **3.87E-11** | # |
| HbA1c% | % | 5.1 | 0.2 | 6 | 0.6 | **5.30E-10** | # |
| Glucose AUC | min*mg/dL | 25272.7 | 2791.2 | 39823.4 | 9380.4 | **3.60E-11** | # |
| Insulin AUC | min*pM | 12663.1 | 5720.4 | 24479.4 | 11337.5 | **1.38E-06** | # |
| C-Pep AUC | min*nM | 317.5 | 111.1 | 480.3 | 145.5 | **3.76E-06** | # |
| GLP1 AUC | min*PM | 1599 | 895.7 | 2563.2 | 1277.7 | **7.59E-04** | # |
| FBG | mg/dL | 89 | 7 | 122 | 20 | **5.89E-12** | # |
| Fasting insulin | pM | 16.1 | 13.6 | 39.7 | 26.8 | **2.89E-05** | # |
| Fasting C-Pep | pM | 267.4 | 156.3 | 1001.2 | 424.6 | **1.48E-12** | # |
| Fasting GLP1 | pM | 5.1 | 3.2 | 8.7 | 4.9 | **7.34E-04** | # |
| HOMA-IR |  | 0.6 | 0.5 | 2 | 1.4 | **2.26E-06** | # |
| Type 2 diabetes | % positive (n) | 0 (0) |  | 52.8 (19) |  | **1.00E-06** | & |
| Prediabetes | % positive (n) | 0 (0) |  | 38.9 (14) |  | **1.00E-05** | & |
| Obesity | % positive (n) | 0 (0) |  | 69.4 (25) |  | **1.00E-08** | & |
| Overweight | % positive (n) | 0 (0) |  | 22.2 (8) |  | **7.54E-03** | & |
| Normal weight | % positive (n) | 100 (28) |  | 8.3 (3) |  | **<1E-10** | & |
| Diabetes medication<br>biguanide (Metformin) | % positive (n) | 0 (0) |  | 63.9 (23) |  | **1.00E-07** | & |
| Diabetes medication<br>biguanide-DPP4i<br>(Metformin-Linagliptin) | % positive (n) | 0 (0) |  | 2.8 (1) |  | 1.00E+00 | & |
| Diabetes medication<br>SGLT2i (Dapagliflozin) | % positive (n) | 0 (0) |  | 2.8 (1) |  | 1.00E+00 | & |
| Diabetes medication<br>other | % positive (n) | 0 (0) |  | 0 (0) |  | 1.00E+00 | & |

**Figure 1:**
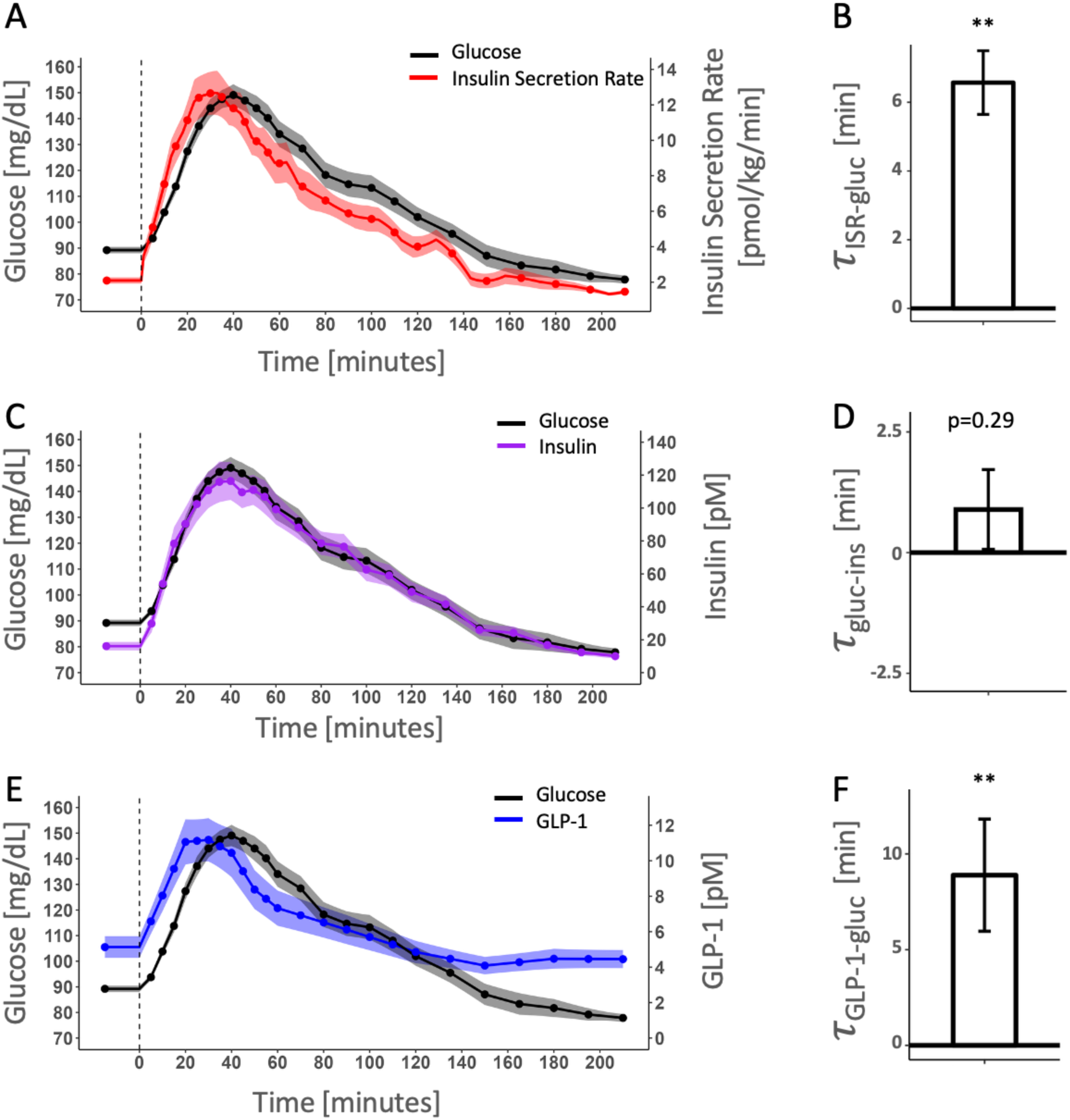
Dynamics of glucose, insulin, insulin secretion rate (ISR), and GLP-1 in healthy participants. A. Average glucose (black) and ISR (red) profiles following ingestion of 50 g glucose. B. Delay between glucose levels and ISR. C. Average glucose (black) and insulin (purple) profiles following ingestion of 50 g glucose. D. Delay between insulin and glucose levels. E. Average glucose (black) and total GLP-1 (blue) profiles following ingestion of 50 g glucose. F. Delay between glucose and total GLP-1 levels. N=28. Error bars in B,D,F, represent the standard error of the mean. Shaded regions in A,C,E represent the standard error of the mean. ** p<0.01 by 1-sample t-test.

To quantify the temporal relationship between these profiles, we performed a cross-correlation analysis, defining *τ_ISR_*_−g*luc*_ as the time shift yielding the maximal correlation coefficient between the ISR and glucose profiles for each participant. We found that *τ_ISR_*_−g*luc*_ = 6±1 minutes (p<0.01; Figure 1A-B, S1B), yielding median maximal correlation of 0.93 across participants. In comparison, the median correlation between ISR and glucose profiles with no time shift was 0.79 (p<0.01, Figure S1C). This analysis demonstrates the anticipatory nature insulin secretion and quantifies the anticipatory time to be 6±1 minutes.

We similarly compared the temporal relationship between the ISR and insulin profiles, and found that the ISR preceded the insulin profile by *τ_ISR_*_−*ins*_ = 6±1minutes (p<0.01; Figure S1D-E) consistent with the 4-6 minutes half-life of insulin in the blood. Since *τ_ISR_*_−g*luc*_ ≈ *τ_ISR_*_−*ins*_, we expected the plasma insulin and blood glucose profiles to overlap. Indeed, we there was no time shift between the glucose and insulin profiles *τ*_g*luc*−*ins*_ = -1±1 minutes, p=0.3; Figure 1C-D, S1B).

We validated these findings by re-analyzing external independent data from Dao *et al*.(30), where healthy participants underwent 25 g and 50 g glucose challenges. Consistent with our findings, ISR was best correlated with future glucose *τ_ISR_*_−g*luc*_=5±1 minutes (50 g, p<0.01) and *τ_ISR_*_−g*luc*_=7±1 minutes (25 g, p<0.01, Fig S1E). Plasma insulin and glucose profiles temporally overlapped in this cohort as well (*τ*_g*luc*−*ins*_ =-2±2 and -1±1 minutes, respectively, Fig S1F).

We analyzed the total GLP-1 profile and found it was best correlated with future glucose profile by *τ_GLP_*_1−g*luc*_ = 9±3 minutes (p<0.01; Figure 1E-F, S1B). In conclusion, in healthy individuals, anticipatory insulin secretion leads to overlapping glucose and insulin profiles.

### Disruption of Anticipatory Insulin Secretion in Patients with Hyperglycemia and Obesity

To assess how metabolic dysfunction affects the temporal relationships between glucose, insulin, ISR, and GLP-1, we recruited 36 participants with varying degrees of BMI and glucose intolerance (mean BMI = 34.6±8.3 kg/m^2^, FBG = 122±20 mg/dL, Table 1). This cohort was older and exhibited elevated fasting blood glucose, HbA1c, and fasting plasma insulin, C-peptide, and GLP-1 concentrations, as well as higher postprandial area under the curves (AUC) for glucose, insulin, and GLP-1. The majority of the cohort were diagnosed with type 2 diabetes (n = 19) or prediabetes (n = 14). Some participants had normoglycemia but met criteria for class III obesity (n = 3). Metformin was the primary treatment for dysglycaemia (Table 1).

The ISR profile in these participants was shifted by *τ_ISR_*_−g*luc*_ = 12±1 minutes compared to the glucose profile (p<0.01; Figure 2A-B, S2A-C), which is longer than in healthy participants (p<0.01; Figure 2B). The delay between plasma insulin and ISR was also extended, *τ_ISR_*_−*ins*_ = 9±1 minutes, p<0.01; Figure S2D), consistent with reduced insulin clearance rates associated with obesity or dysglycaemia(14, 16, 31, 32). Consequently, there was no temporal lag between plasma insulin and glucose (*τ*_g*luc*−*ins*_=-2±2;p=0.4, Figure 2C-D, S2A). Finally, we note that the GLP-1 profile was shifted by *τ_GLP_*_1−g*luc*_ = 28±3 min (p<0.01; Figure 2E-F, S2A), much longer than in healthy participants, reflecting the late decline in blood glucose levels in individuals with obesity or dysglycaemia.

**Figure 2:**
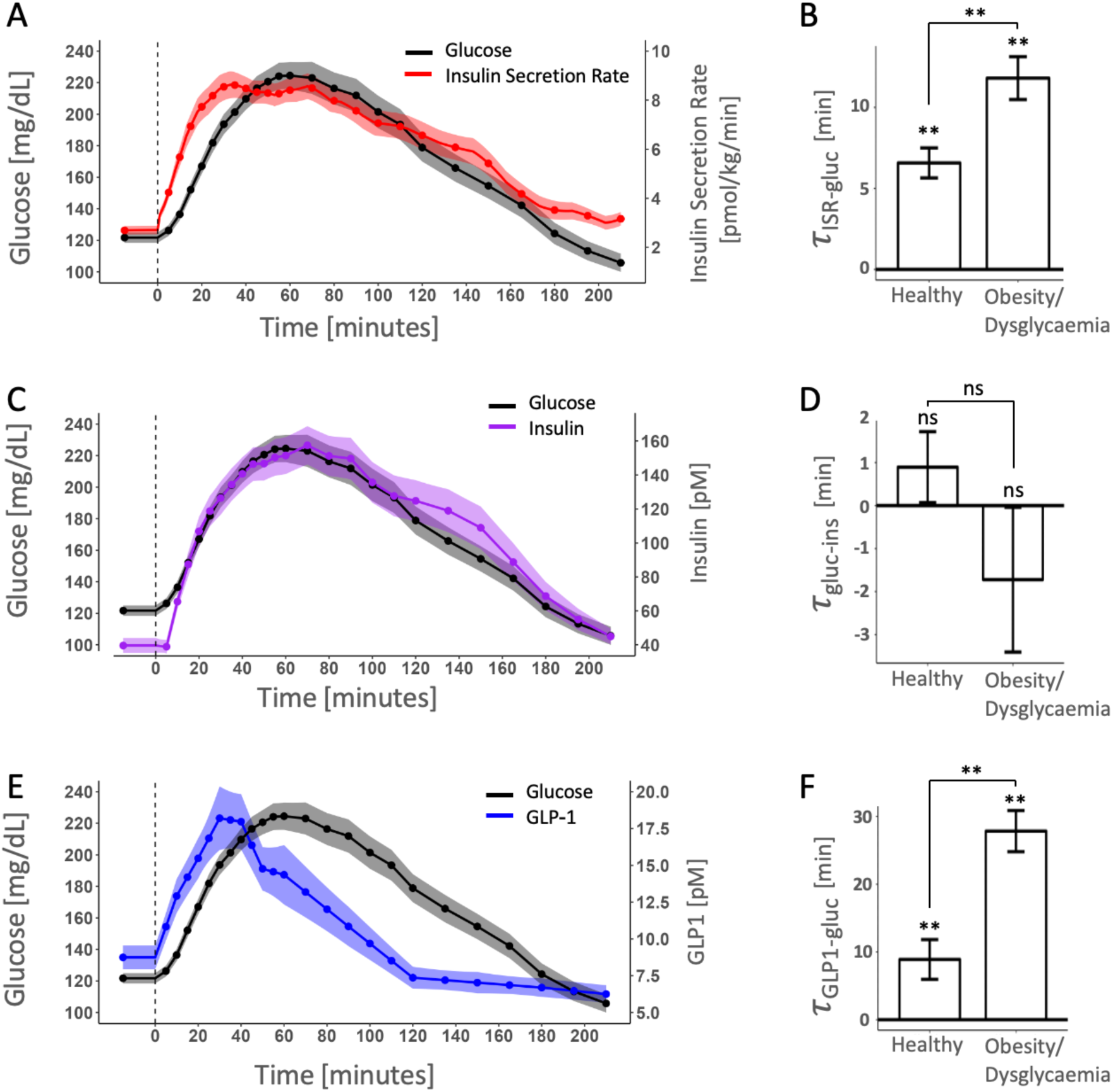
Dynamics of glucose, insulin, insulin secretion rate (ISR), and GLP-1 in participants with obesity and/or dysglycaemia. A. Average glucose (black) and ISR (red) profiles following ingestion of 50 g glucose. B. Delay between glucose levels and ISR in healthy participants and participants with obesity/dysglycaemia. C. Average glucose (black) and insulin (purple) profiles following ingestion of 50 g glucose. D. Delay between insulin and glucose levels in healthy participants and participants with obesity/dysglycaemia. E. Average glucose (black) and total GLP-1 (blue) profiles following ingestion of 50 g glucose. F. Delay between glucose and total GLP-1 levels in healthy participants or participants with obesity/dysglycaemia. N=36. Error bars in B,D,F, represent the standard error of the mean. Shaded regions in A,C,E represent the standard error of the mean. ** p<0.01 by 1-sample t-test for each condition and 2-sample t-test when comparing between healthy participants and participants with obesity/dysglycaemia.

### Disruption in Anticipatory Insulin Secretion is Associated with High Glucose AUC

The extent of anticipatory time shift between ISR and glucose, *τ_ISR_*_−g*luc*_, was most highly correlated with the glucose AUC (ρ=0.43, p<0.01, Figure 3A,B, S3A). The longer temporal shift between ISR and glucose profiles, implies that insulin secretion declines prematurely in participants with postprandial hyperglycemia. Since GLP-1 acts as a feed-forward signal, we suspected that this disruption of anticipatory ISR is related to the wider temporal shift between the GLP-1 and glucose profiles: GLP-1 levels fall 28±3 minutes before glucose levels decline, compared to 9±3 minutes in healthy individuals. In support, we identified a positive correlation between time shift between GLP-1 and glucose (*τ_GLP_*_1−g*luc*_) and *τ_ISR_*_−g*luc*_ (ρ=0.41, p<0.01) (Figure 3C). There was also a significant correlation between *τ_GLP_*_1−g*luc*_ and glucose AUC (ρ=0.50, Figure 3D). In conclusion, slower dynamics of glucose together with rapid dynamics of GLP-1 may disrupt fine-tuned anticipatory insulin secretion, resulting in pre-mature reduction in ISR.

**Figure 3.**
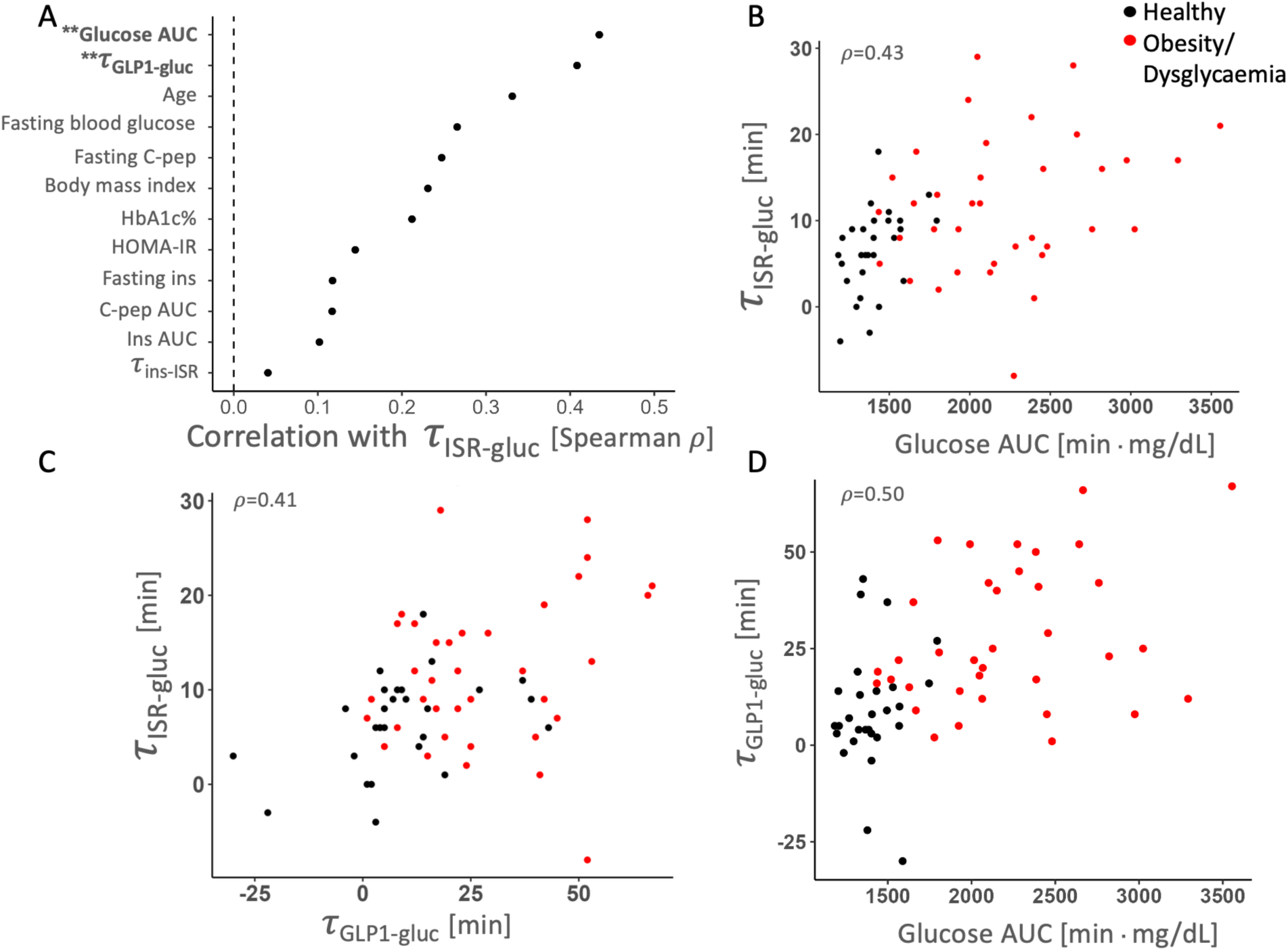
Premature decline in ISR and GLP-1 is associated with higher glucose area under the curve A. Spearman correlation coefficient between the glucose-ISR delay and the main characteristics in all participants (N=64). Factors in bold and ** show a statistically significant correlation. B. Scatterplot showing positive correlation in the delay between the glucose and ISR profiles, and the glucose area under the curve. C. Scatterplot showing positive correlation in the delay between the glucose and GLP-1 profiles, and the delay between the glucose and ISR profile. D. Scatterplot showing positive correlation in the delay between the glucose and GLP-1 profiles and the glucose area under the curve. Black dots in B,C,D mark healthy participants, red dots mark participants with obesity and/or dysglycaemia.

### Semaglutide treatment abolishes the temporal shift between ISR and glucose

According to the model presented above, GLP-1 is part of the feed-forward signal controlling anticipatory insulin secretion. We hypothesized that constitutive activation of the GLP-1 receptor would increase ISR, but might mask the endogenous feed-forward signal, leading to a loss of anticipatory insulin secretion altogether. A sub-cohort of the participants with obesity and/or dysglycaemia commenced treatment with semaglutide (n=14, baseline BMI=38.9±11.0 kg/m^2^, FBG=116±12 mg/dL, Table 2). Average treatment duration was 6±1 months and treatment dose was 1-2.4 mg/week.

**Table 2:**
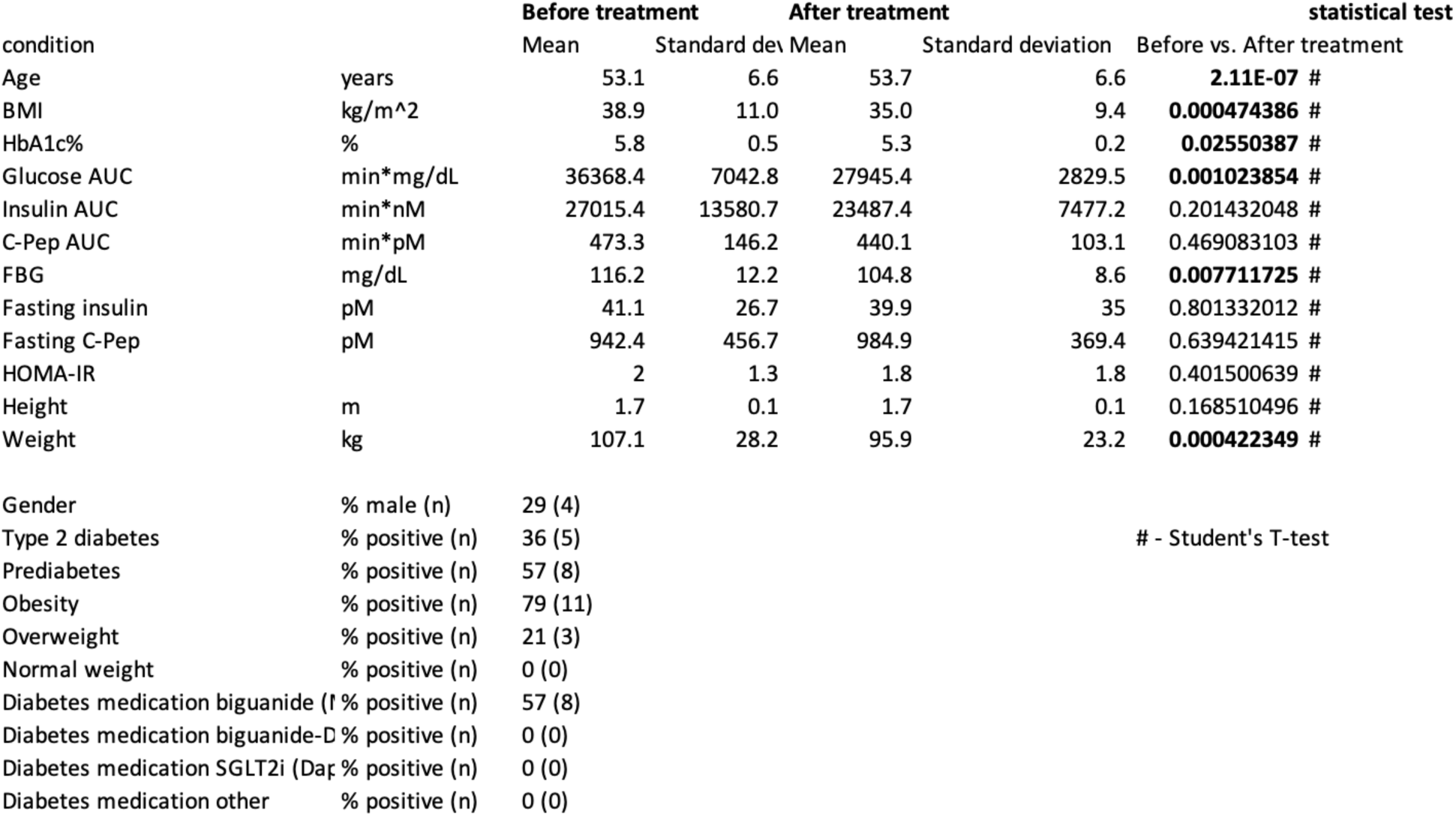
characteristics of participants with obesity/dysglycaemia that received semaglutide before and after treatment.

Post-treatment, participants exhibited significant weight loss (mean BMI reduction 3.7±2.4 kg/m^2^), improved glycemic markers (Table 2), and increased maximal ISR to levels of healthy participants (Figure S4A-B). In agreement with our hypothesis, semaglutide treatment abolished the time shift between ISR and glucose: *τ_ISR_*_−g*luc*_= 2±2 minutes, p=0.4; Figure 4A-B, S4C), compared to a 12±2 minutes time shift before treatment in the same individuals (p<0.01 Figure 4B). The delay between ISR and plasma insulin persisted *τ_ISR_*_−*ins*_ = 11± 2 minutes, p<0.01, Figure S4D-E), and as a result, the plasma insulin profile which overlapped before treatment, now significantly lagged behind the glucose profile *τ*_g*luc*−*ins*_ = 10±3 minutes, p<0.01; Figure 4C-D, S4C). The endogenous GLP-1 profiles could not be quantified due to an apparent assay cross-reactivity with semaglutide (Figure S4F).

**Figure 4.**
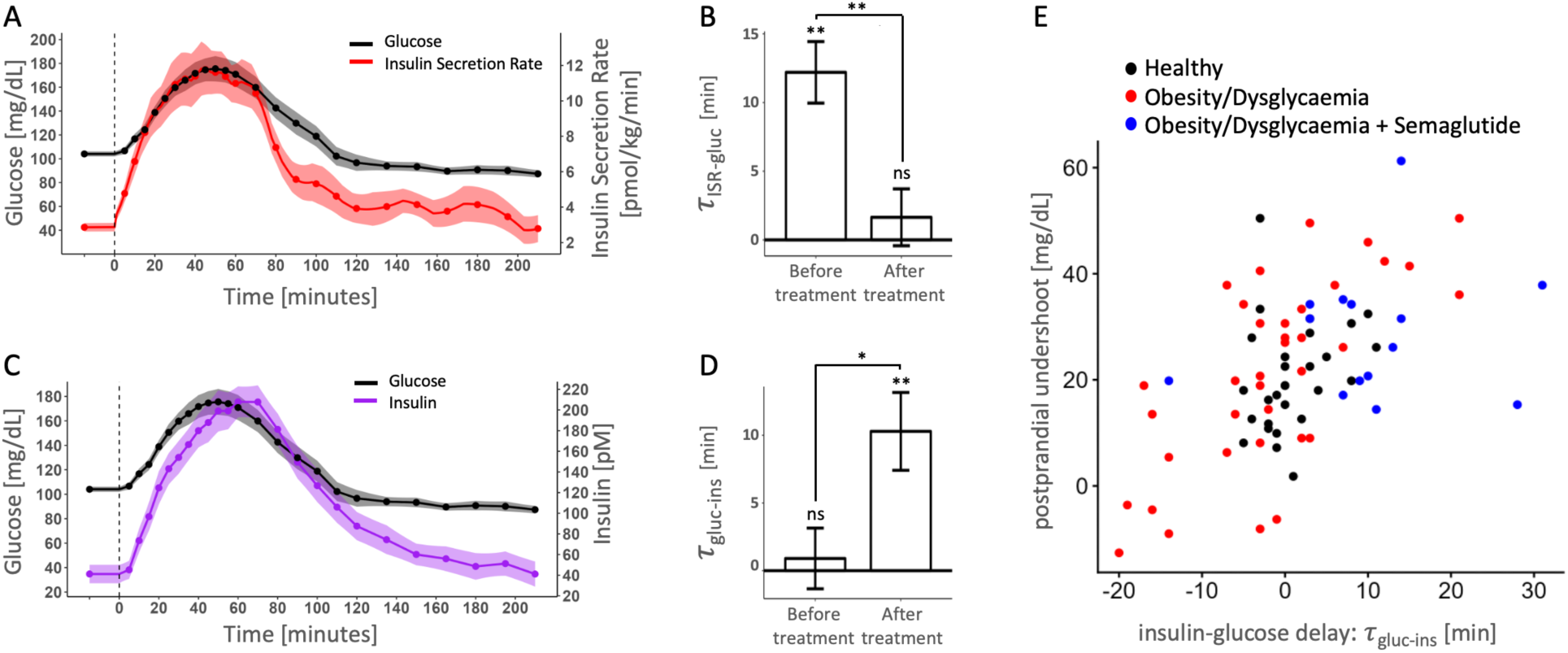
Dynamics of glucose, insulin, and insulin secretion rate (ISR) in participants with obesity and/or dysglycaemia after treatment with semaglutide. A. Average glucose (black) and ISR (red) profiles following ingestion of 50 g glucose. B. Delay between glucose levels and ISR before and after treatment with semaglutide. C. Average glucose (black) and insulin (purple) profiles following ingestion of 50 g glucose. D. Delay between insulin and glucose levels before and after treatment with semaglutide. E. Scatterplot showing positive correlation between the time delay of insulin and glucose and the postprandial undershoot in glucose levels. Black dots mark healthy participants, red dots mark participants with obesity and/or dysglycaemia, and blue dots mark participants after treatment with semaglutide. N=14 in A-D. Error bars in B,D, represent the standard error of the mean. Shaded regions in A,C represent the standard error of the mean. *,** p<0.05, p<0.01 for each condition by 1-sample t-test and between conditions by paired t-test..

A lag between insulin and glucose implies that insulin levels remain high postprandially whereas glucose levels start to decrease, potentially driving a postprandial undershoot of glucose levels. We defined this undershoot as the difference between the participant’s fasting glucose and minimal postprandial glucose. The postprandial glycemic undershoot correlated strongly with the magnitude of the insulin-glucose delay (ρ=0.78, p<0.01; Figure 4E), and was larger in individuals treated with semaglutide (Figure S4G)

## Discussion

In this study, we characterized the temporal dynamics of glucose, insulin, ISR, and GLP-1 in healthy individuals, individuals with obesity, pre-diabetes and type 2 diabetes, and a subset of these patients treated with semaglutide. In healthy participants, the ISR profile correlates with the glucose profile in the near future (6±1 minutes ahead), demonstrating and quantifying anticipatory insulin secretion. These quantitative findings were validated *post-hoc* by external data from Dao *et al*.(30).

Anticipatory secretion was disrupted in patients with obesity and/or dysglycaemia. ISR preceded the glucose profile by 12±2 minutes, twice as long as in healthy individuals. This large gap reflects a premature reduction in ISR, contributing to a higher glucose AUC. GLP-1 signaling provides some of the information required for anticipatory insulin secretion via a feed-forward mechanism. GLP-1 preceded glucose by 9±3 minutes in healthy individuals. However, in patients with obesity and/or dysglycaemia, GLP-1 preceded glucose by 28±3 minutes. This large gap reflects the slower dynamics of glucose clearance, peaking 60 minutes after glucose consumption, while the GLP-1 dynamics remain rapid, peaking 30 minutes after ingestion. The discordance between the GLP-1 and glucose dynamics may underlie the premature decrease in ISR, contributing to hyperglycemia. Obesity and dysglycaemia are associated with slow insulin clearance rate(33), which increase the lag between ISR and plasma insulin. The early decline in ISR together with longer clearance rate coincidently resulted in relative synchronization of plasma glucose and insulin.

Semaglutide treatment leads to a constitutive GLP1R activation. Semaglutide treatment improved weight and glycemia and increased the maximal ISR, reaching values of lean individuals. However, semaglutide treatment may have saturated endogenous GLP-1 signaling, leading to loss of temporal information provided by the gut. The compensated feed-forward signal abolished anticipatory insulin secretion, resulting in a delay between insulin and glucose profiles.

What is the function of anticipatory insulin secretion, and the outcome of its loss? We hypothesize the anticipatory control balances between two demands: minimizing postprandial hyperglycemia while avoiding postprandial glycemic undershoots. The rapid initial rise in ISR, corresponding to the rising glucose levels, curbs postprandial hyperglycemia. This can be directly observed in our data: premature decline in ISR, corresponding to a 12±2 minutes difference between the ISR and glucose profiles, correlates with larger glucose AUC. An early decline in ISR, preceding glucose drop by 5-10 minutes in healthy individuals, prevents a postprandial undershoot in glycemia. Indeed, loss of anticipatory insulin secretion which occurs in semaglutide treated patients is associated with a lag between insulin and glucose, and larger postprandial glucose undershoots.

### GLP-1 provides temporal information for glucose regulation

Mechanistically, incretin hormones (GLP-1 and GIP) induce satiety, reduce gastric emptying, and amplify glucose-dependent insulin secretion(2, 18, 20, 34). Their early rise contributes to the rapid rise of ISR, establishing the early temporal lead of ISR over glucose. The decline in GLP-1, and potentially other gut-derived signals, results in a reduction in ISR that precedes the decline in blood glucose levels, reducing the risk of large postprandial glucose dips.

Anticipatory regulation of glucose was reported and incorporated in models for glucose-insulin dynamics(16, 35–43). GLP-1, acting as a feed-forward signal that amplifies ISR in response to glucose, can explain quantitatively the time shift between ISR and glucose profiles. Mathematical analysis suggests *τ_ISR_*_−g*luc*_ ≈ 0.5 · *τ_GLP_*_1−g*luc*_(Supp. Information), as we observe in our data (Figure S5A-E). In patients with obesity and dysglycaemia, the gap between the GLP-1 and glucose profiles was 28±3 minutes. This may have contributed to the 12±2 minutes difference between the ISR and glucose profiles.

GLP-1 is one of several factors, along with GIP, glucagon, intra-islet paracrine signaling, neural signals and intracellular factors, that modulate ISR(1–6, 44, 45). Neuronal signaling and incretins can convey temporal information on future glucose levels. We hypothesized that constitutive activation of the GLP-1 receptor via semaglutide would saturate an endogenous temporal signal, blinding the β cells to temporal cues. In this scenario, secretion would be driven solely by current glucose levels, as we observed in patients treated with semaglutide. Semaglutide improved clinical parameters in treated participants but abolished the tight temporal coordination between insulin and glucose levels observed in non-treated individuals. Treatments that augment endogenous GLP-1 signaling without affecting their dynamics may restore anticipatory insulin secretion.

### Control Theory Perspective

An ideal homeostatic system minimizes the phase lag between the controlled variable and the controller’s output(22, 23). In this case: blood glucose and plasma insulin levels. The kinetics of insulin clearance introduces an inherent delay between insulin secretion rate and circulating insulin levels. If the levels of glucose decline rapidly enough, such a delay can create an overshoot in the control signal, leading to a postprandial dip, or undershoot in glucose levels(45–47).

A feed-forward signal can overcome this pitfall(48). By utilizing feed-forward control, the controller responds to the future level of the controlled variable. The feed-forward signal and phase lag cancel each other out, resulting in a temporal overlap between the controlled variable and the controller’s output. GLP-1 fits as a feed-forward signal in this control architecture: it correlates with future levels of glucose, *τ_GLP_*_1−g*luc*_ correlates with *τ_ISR_*_−g*luc*_, and constitutive GLP-1 activation saturates the feed-forward signal, resulting in an overlap between ISR and glucose levels and a postprandial glucose undershoot. Feed forward control is prevalent in engineering and in physiology, affecting gastric and pancreatic exocrine secretion, gut motility, and feeding behaviour(49–51).

Incretins amplify insulin secretion in response to ingested glucose and other macronutrients. How is amplification of insulin secretion compatible with a feed-forward signal? Mathematical analysis shows that dynamically changing the gain signal in the control over ISR, the amplification of insulin secretion is an implementation of feed-forward control overcoming an inherent lag between blood glucose and plasma insulin levels (Supp. Information).

## Limitations

This study has several limitations. The data reflect responses to a liquid glucose challenge, which is different than a response to a meal. The study was not designed to detect the cephalic phase of insulin secretion, which occurs before nutrient absorption(43, 52). We measured total rather than active GLP-1(44); while their dynamics are generally similar, the specific bioactive signal reaching β cells may differ. ISR was derived using ISEC(29, 53), a standard deconvolution method based on population-average C-peptide clearance parameters. Finally, venous sampling yields slightly lower and delayed glucose values compared to arterial blood; we partially mitigated this by warming the drawing site to “arterialize” venous blood(54).

## Conclusions

We demonstrate the anticipatory secretion of insulin in healthy individuals, showing that ISR is correlated with the future glucose profiles by approximately 6 minutes, leading to synchronization between blood glucose and plasma insulin profiles. Anticipatory insulin secretion is disrupted in obesity and dysglycaemia, wherein ISR starts dropping prematurely, potentially contributing to post-prandial hyperglycemia. Our analysis suggests that by amplifying ISR, GLP-1 and potentially other gut-derived signals feed-forward information to the pancreas on future glucose levels. In support, constitutive activation of the GLP-1 receptor by semaglutide disrupts the feed-forward signal. As a result, anticipatory insulin secretion is abolished, and insulin secretion rate tracks current glucose levels. Our findings illuminate and quantify anticipatory insulin secretion, highlight the role of GLP-1 as a feed-forward signal, and demonstrate the disruption of anticipatory insulin secretion in obesity, glucose intolerance and hyperglycemia and following treatment with GLP-1 receptor agonists.

## Data Availability

All data produced in the present study are available upon reasonable request to the authors

## Acknowledgement

We thank members of our group for fruitful discussions and comments on this manuscript.

