## supporting information for "Disruption of Anticipatory Insulin Secretion in Patients with Hyperglycemia and Obesity"

Doron Kleiman[1], Itia Magenheim-Samuel[1], Sarah Knapp-Goldin[1], Sienna Walsh[2], Renee Richens[2], Krisztina Toth[3], Jennifer Evans[3], Giang M Dao[4], Clinton R Bruce[4], Christopher S Shaw[4], Andrew C Betik[4], Vicky Kuriel[4], Shlomi Kotler[5], Greg M Kowalski[4], Tania Markovic[6], Malgorzata Brzozowska[7], Jerry R Greenfield[3,8,9], Rachel Ben-Haroush Schyr[1], Dorit Samocha-Bonet[2,8,10]*, Danny Ben-Zvi[1,11*]

[1] Department of Developmental Biology and Cancer Research, Institute of Medical Research Israel-Canada, the Faculty of Medicine-Hadassah Medical School, the Hebrew University of Jerusalem, Jerusalem
[2] Garvan Institute of Medical Research, Sydney, Australia
[3] The Victor Chang Cardiac Research Institute, Sydney, NSW, Australia
[4] Institute for Physical Activity and Nutrition, Metabolic Research Unit, School of Medicine, Deakin University, Geelong, Victoria, Australia
[5] The Racah Institute of Physics, Hebrew University of Jerusalem, Jerusalem, Israel
[6] Charles Perkins Centre and Faculty of Medicine and Health, The University of Sydney, Sydney, NSW, Australia
[7] St George and Sutherland Campus, School of Clinical Medicine, UNSW, Sydney, NSW, Australia
[8] St. Vincent's Clinical Campus, School of Clinical Medicine, UNSW Sydney, NSW, Australia
[9] Department of Endocrinology and Diabetes, St Vincent's Hospital, Sydney, NSW, Australia
[10] School of Health Sciences, UNSW Medicine and Health, UNSW Sydney, NSW, Australia
[11] Center for Computational Medicine, Faculty of Medicine-Hadassah Medical School, the Hebrew University of Jerusalem, Jerusalem, Israel

#### Extended Methods

The study was approved by the St Vincent's Hospital Human Research Ethics Committee, Sydney, Australia (approval numbers 2019/ETH12216 and 2020/ETH02405). All participants provided written informed consent before the commencement of any study-related procedures.

Participants: Healthy participants were men and women aged 18-60 years who had a normal weight (BMI ≤25 kg/m²), HbA1c ≤5.5 %, not treated with glucose-lowering medications, never smoked, with liver and kidney function tests in the normal range, without any known medical condition.

Participants with obesity/dysglycemia were individuals who had been advised by their managing physician to commence semaglutide treatment for an underlying condition. These included people living with obesity (BMI ≥ 30 kg/m²), with or without dysglycaemia, as well as people without obesity who had prediabetes or type 2 diabetes. Initiating treatment with semaglutide was not mandatory, and not all participants have initiated semaglutide during the study period.

Exclusion criteria relevant to all participants were pregnancy or lactation. In addition, exclusion criteria for the candidates for semaglutide treatment were serum alanine aminotransferase [ALT] and/or aspartate aminotransferase [AST]>3-times the upper limit of normal, estimated glomerular filtration rate eGFR<45 mL/min/1.73 m², haemoglobin <115 g/L (all assessed within the preceding 6 months), type 1 diabetes, active autoimmune disease, malignancy within the previous 3 years, cardiovascular event within the preceding 6 months, prior bariatric, pancreatic, or upper gastrointestinal surgery, history of pancreatitis, normoglycaemia, HbA1c >9.0% (75 mmol/mol), and treatment with insulin, systemic corticosteroids, immunosuppressive agents, or antiviral medications.

##### Study procedures

Screening

Participants attended a screening visit at the Clinical Research Facility at the Garvan Institute of Medical Research. Anthropometric measurements were obtained, and a medical history was recorded. Blood was drawn and sent to a commercial laboratory to assess eligibility.

Study visit

Participants attended the Clinical Research Facility following a 10-12-hour overnight fast to undergo a modified glucose challenge test. In the 48 hours preceding the study visit, participants were instructed to avoid strenuous physical activity and alcohol consumption. For the 3 days prior to the study, participants were advised to consume three main meals per day (snacks permitted) and to include at least one serving of carbohydrate with each meal.

On arrival, weight, height, waist and hip circumference, and blood pressure were measured. An intravenous cannula was inserted into the antecubital vein, and the cannulated arm was kept warm using a heating blanket. Two baseline blood samples were collected 15 minutes apart. Participants then consumed a 50 g glucose solution (200 mL) within 5 minutes, followed by serial blood sampling for 3.5 hours (every 5 minutes during the first hour, every 10 minutes during the second hour, and every 15 minutes for the remainder of the study period).

Participants treated with a glucose-lowering medication at baseline underwent a medication wash-out period based on four-times the elimination half-life of the respective drug. If not treated with metformin, participants were transitioned to metformin, which was discontinued 24 hours before the baseline study visit.

Participants who commenced treatment with semaglutide, underwent an identical study protocol following treatment (aiming for 5 - 7 months of continuous treatment, 2.4mg/weak). Study staff have maintained frequent contact with these participants via telephone follow-up throughout the treatment period.

**Biochemical analyses**

Whole blood glucose concentrations were measured using the Nova StatStrip glucose monitoring system (Nova Biomedical, USA). Blood samples were centrifuged, and plasma was aliquoted and stored at −70 °C until analysis. Plasma insulin and C-peptide concentrations (27 samples per person) were measured using enzyme immunoassays (#90095 and #80954; Crystal Chem, USA). For the measurement of GLP-1, blood samples were collected into P800 tubes (BD Vacutainer®, Becton, Dickinson and Company, USA), and the plasma stored at −70 °C until analysis. Plasma total GLP-1 concentrations were determined (11 samples per person) using the V-Plex Total GLP-1 kit (K1503PD-2; Meso Scale Discovery, USA).

**Calculations**

C-peptide concentrations were used to calculate ISR using the ISEC deconvolution software program[30]. HOMA-IR was calculated: Fasting glucose (mg/dL) x Fasting insulin (mU/L)/405.

Cross correlations between glucose, insulin, ISR, and GLP-1 were calculated by custom software in R.

**Statistical Analysis**

Spearman's correlation coefficient was calculated determining the correlation throughout the text. A 1-sample Studnet'st-test was used to determine if a specific delay, was different than zero. A 2-sample Student's t-test, Mann-Whiteny U-test or 1-way ANOVA with Tukey post-hoc test were was used to compare parameters in healthy participants and participants with obesity and/or dysglycaemia and following treatment with semaglutide. See specific details in figure captions.

#### Supporting Figures

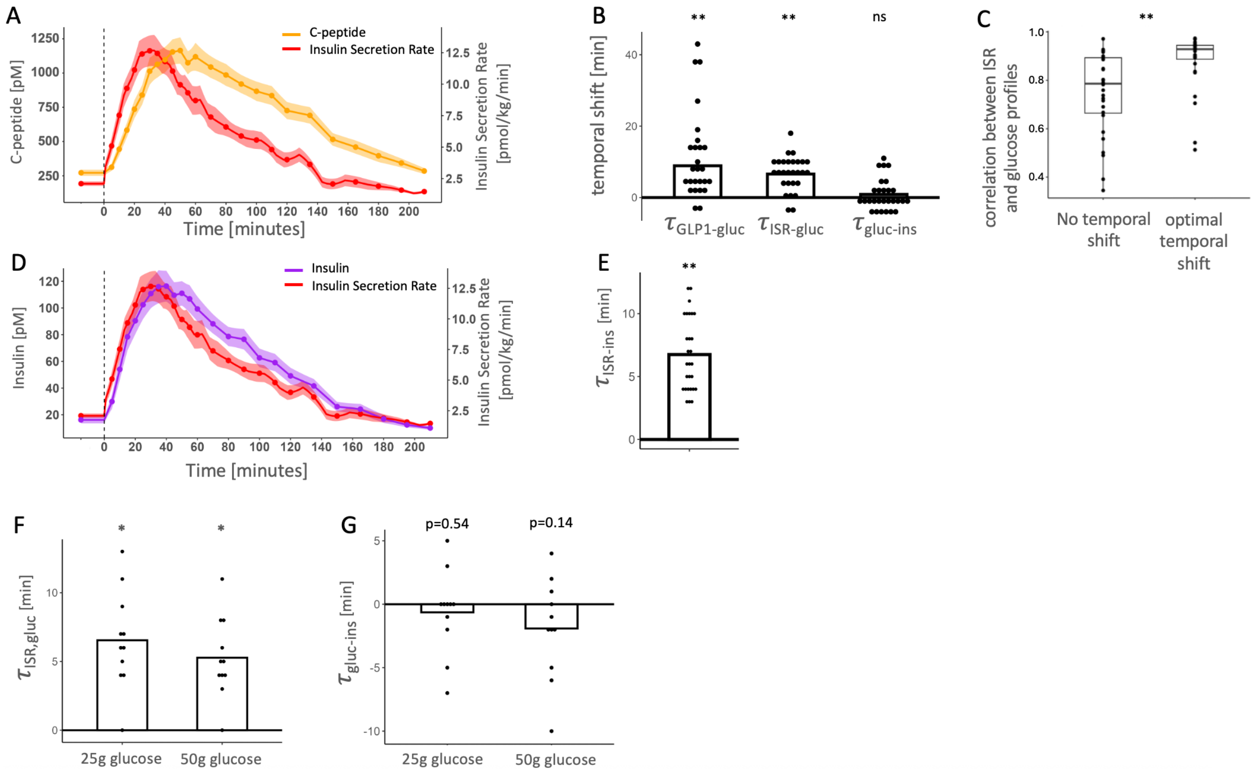


Figure S1: Dynamics of glucose, insulin, insulin secretion rate (ISR) and GLP-1 in healthy participants.

A. Average C-peptide (yellow) and ISR (red) profiles following ingestion of 50 g glucose. Data refer to Figure 1.

B. Glucose GLP-1 delay; Glucose-ISR delay and insulin-glucose delay. Data refer to Figure 1.

C. Spearman correlation between glucose and ISR profiles with no temporal shift (left) and in the temporal shift where maximal correlation in observed (right).

D. Average ISR (red) and insulin (purple) profiles following ingestion of 50 g glucose. Data refer to Figure 1.

E. Delay between insulin and ISR. Data refer to Figure 1.

F. Delay between glucose and ISR in data from Dao et al, 2025[2], in 11 participants who ingested 50 or 25 g of glucose.

G. Delay between insulin and glucose in data from Dao et al, 2025[2], in 11 participants who ingested 50 or 25 g of glucose.

N=28 in A-D. Shaded regions in (A,D,F) represent the standard error of the mean. *p<0.05, ** p<0.01 ns non-significant by 1-sample t-test in (B,E,F,G) and by Mann-Whitney U-test in (C).

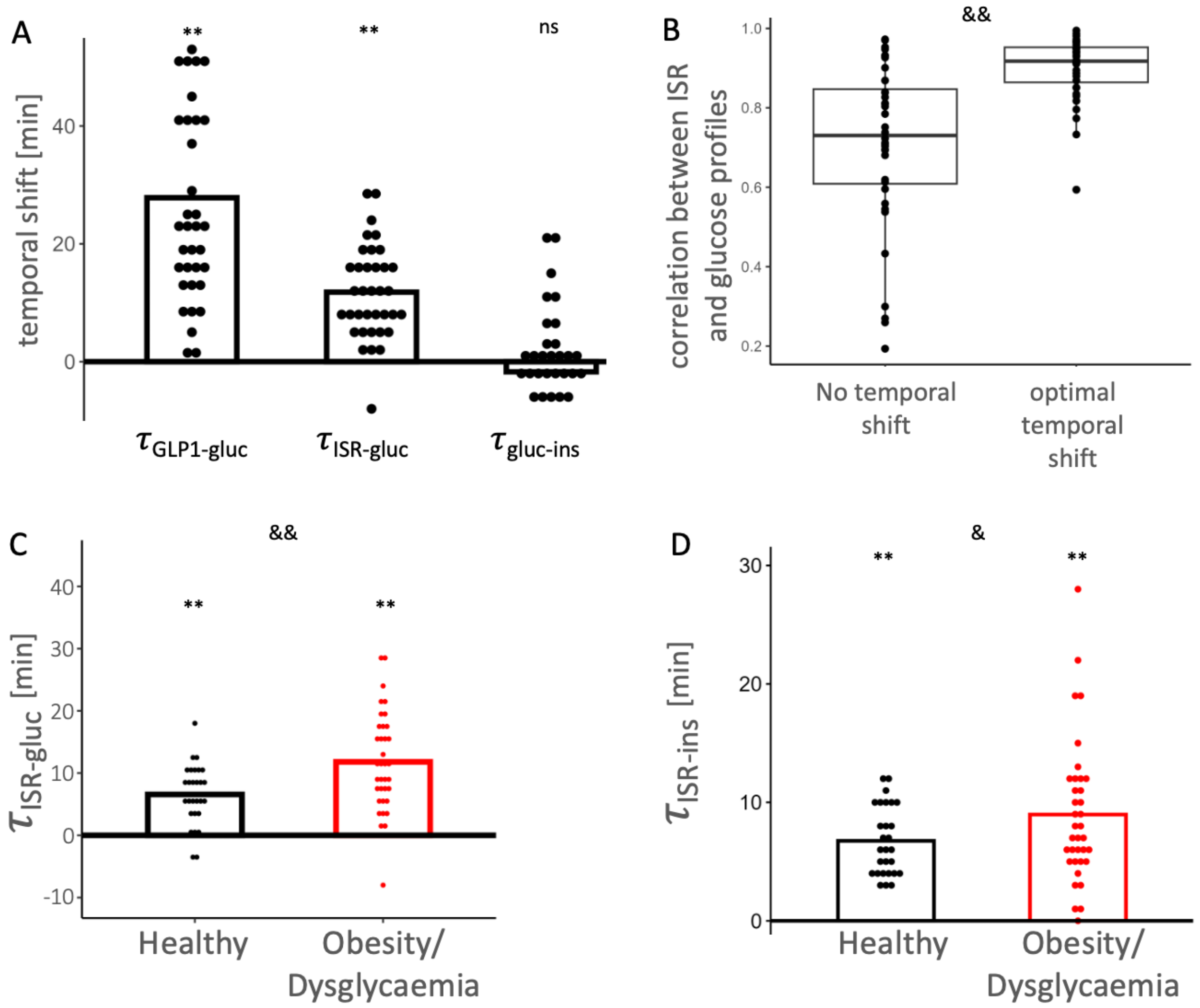


Figure S2: Dynamics of glucose, insulin, insulin secretion rate (ISR), and GLP-1 in participants with obesity and/or dysglycaemia.

A. Glucose GLP-1 delay; Glucose-ISR delay and insulin-glucose delay. Data refer to Figure 2.

B. Spearman correlation between glucose and ISR profiles with no temporal shift (left) and in the temporal shift where maximal correlation in observed (right).

C. Delay between glucose and ISR in healthy participants (black) and in participants with obesity and/or dysglycaemia (red)

D. Delay between insulin and ISR in healthy participants (black) and in participants with obesity and/or dysglycaemia (red)

** p<0.01 by 1-sample t-test (A,C,D). [&, &&] p<0.05, 0,01 comparing two groups in Mann Whitney U-test (B) and Student's t-test in (C,D).

A

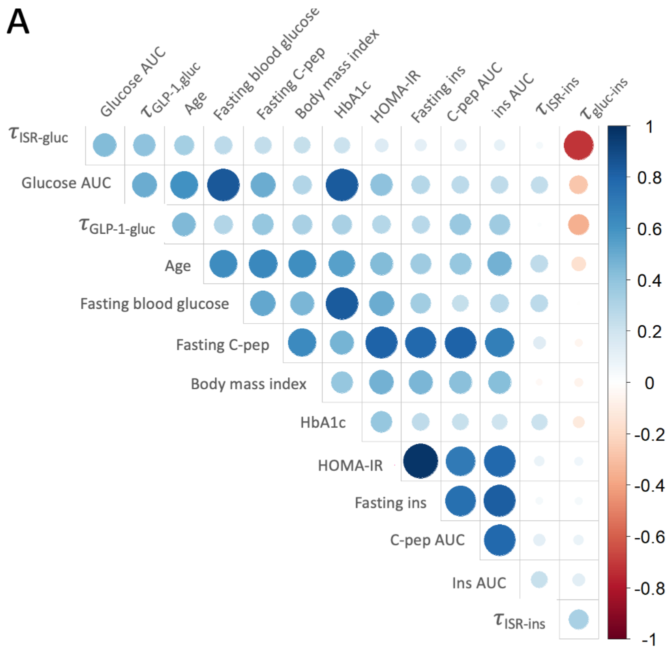


Figure S3. GLP1 acts as a feed forward signal for insulin secretion rate

A. Spearman correlation coefficients between the main glycemic and anthropometric characteristics of the study participants.

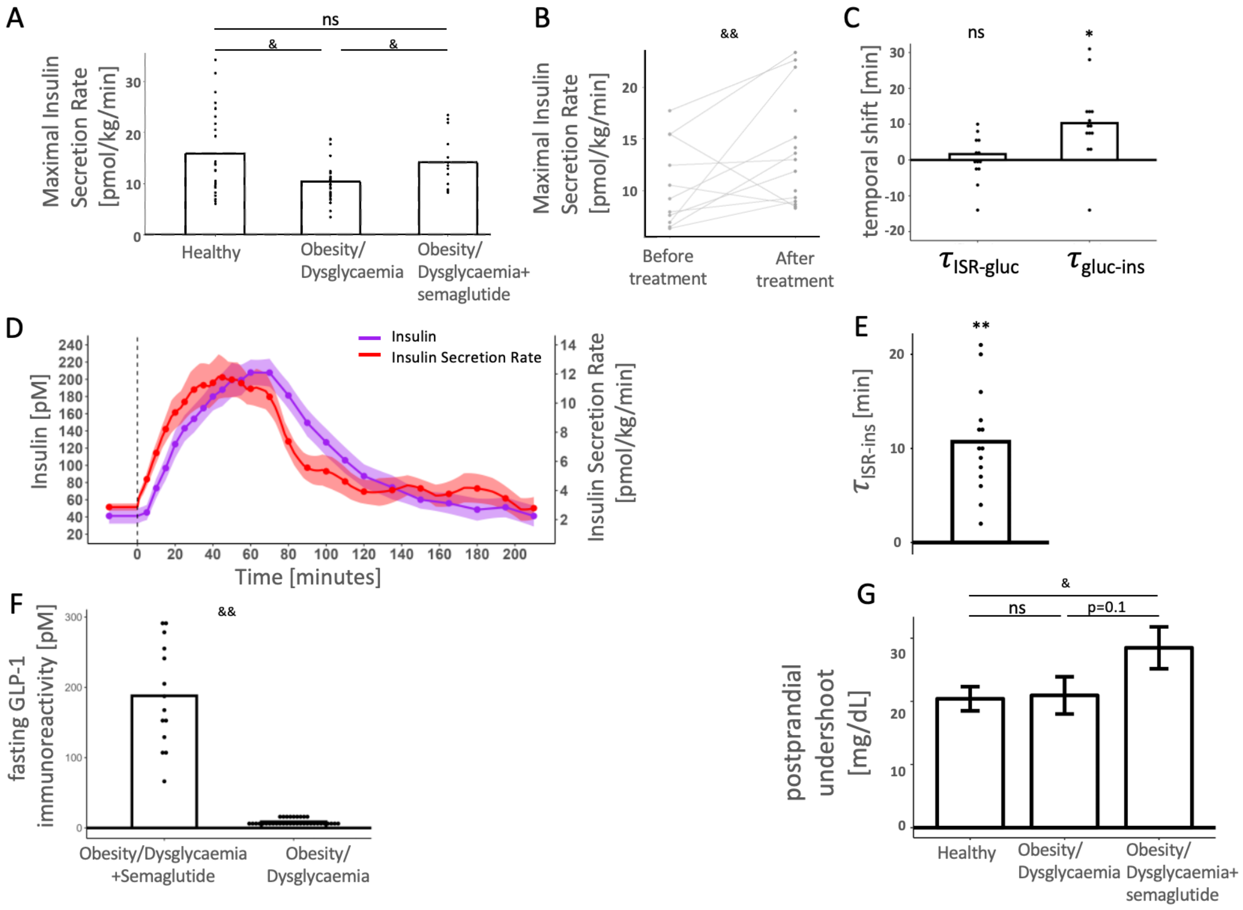


Figure S4: Dynamics of glucose, insulin, and insulin secretion rate (ISR) in participants with obesity and/or dysglycemia after treatment with semaglutide.

A. Maximal ISR in healthy participants, participants with obesity/dysglycaemia, and those threated with semaglutide.

B. Maximal ISR in participants with obesity/dysglycaemia before and after treatment with semaglutide.

C. Glucose-ISR delay and insulin-glucose delay. Data refer to Figure 4.

D. Average ISR (red) and insulin (purple) profiles following ingestion of 50 g glucose. Data refer to Figure 4.

E. Delay between insulin and ISR. Data refers to Figure 4

F. Fasting GLP-1 immunoreactivity in participants with obesity and/or dysglycaemia treated with semaglutide, and in participants not treated with semaglutide, demonstrating cross-reactivity between semaglutide and the GLP-1 immunoassay.

G. Difference between minimal postprandial glucose and fasting blood glucose in healthy participants, participants with obesity/dysglycaemia, and those threated with semaglutide.

*, ** in C,E p<0.05, 0.01 by 1-sample Student's t-test. [&,&&] p<0.05,0.01 if Tukey post-hoc test following one way ANOVA in A,G, paired t-test in B, and 2-sample Student's t-test in F.

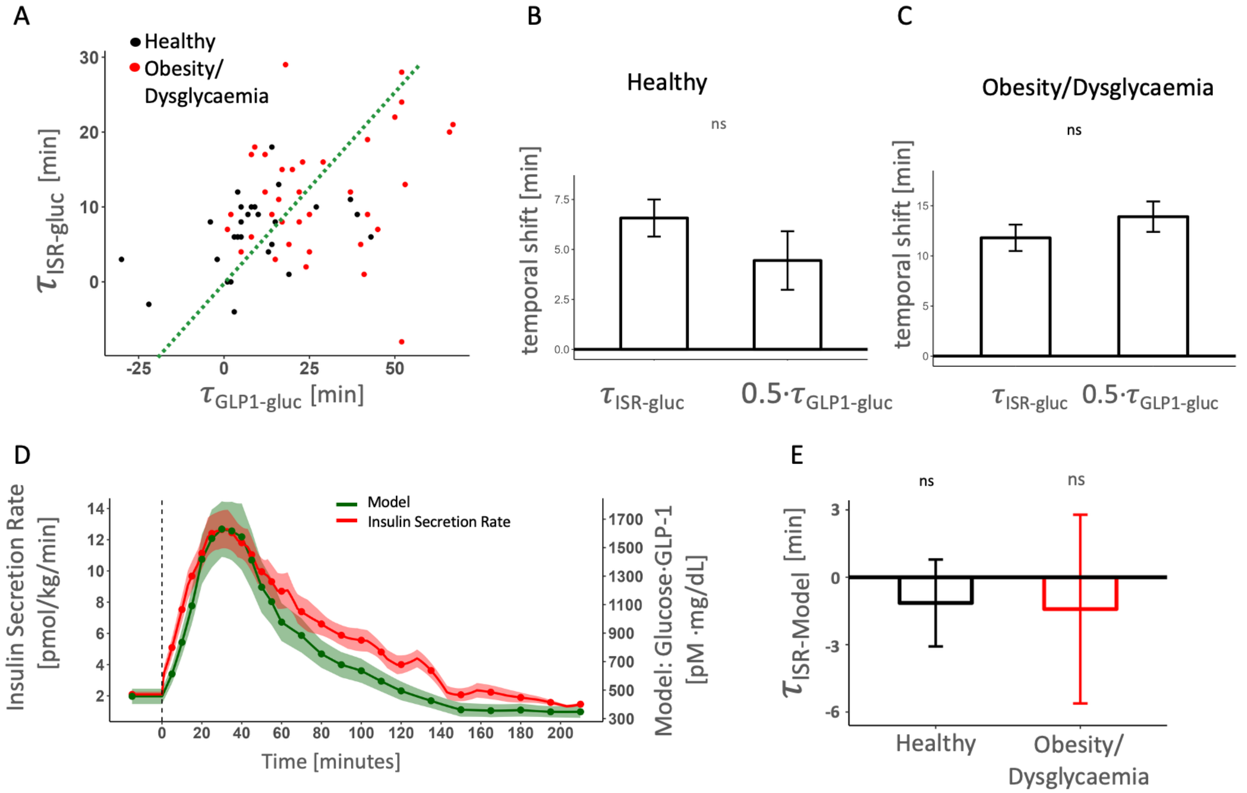


Figure S5: mathematical modelling of anticipatory insulin secretion. As a function of glucose and GLP-1 profiles.

A. Scatterplot showing positive correlation in the delay between the glucose and GLP-1 profiles, and the delay between the glucose and ISR profile. The green dotted line marks the theoretical model, $\tau_{ISR-gluc} \approx 0.5 \cdot \tau_{GLP1-gluc}$.

B. Temporal shift between ISR and glucose $\tau_{ISR-gluc}$, and one half Glp1 and glucose $\frac{\tau_{GLP1-gluc}}{2}$ in healthy participants.

C. Temporal shift between ISR and glucose $\tau_{ISR-gluc}$, and one half Glp1 and glucose $\frac{\tau_{GLP1-gluc}}{2}$ in participants with obesity.

D. Measured ISR profile (red) and model prediction (green) in healthy participants.

E. The Delay between model and measured ISR, showing no difference in healthy participants and patients with obesity and/r dysglycaemia.

differences between $\tau_{ISR-gluc}$ and $\frac{\tau_{GLP1-gluc}}{2}$ non-significant by 2-sample Student's t-test (B,C). Error bars in B,C,E represent standard error of the mean. Shaded areas in D denote standard error. ns by 1 sample t-test in E.

**Mathematical analyses**

### 1 Introduction

Here we show that a convolution of a wide pulse, such as the postprandial insulin secretion rate profile which lasts about 100 minutes, convoluted with a fast decaying kernel, such as the insulin clearance profile which has a time scale of 5-10 mintues, results in a function which is similar to the wide pulse (insulin secretion rate) shifted by the characteristic time of the fast decaying Kernel.

Let $f(t)$ be a wide, smooth pulse and $g(t)$ be a sharply declining, causal kernel. We define the properties of $g(t)$ by its zeroth and first moments:

$$\int_{-\infty}^{\infty} g(x)\,dx = G \quad \text{(Gain)} \tag{1}$$

$$\int_{-\infty}^{\infty} xg(x)\,dx = G \cdot \tau \quad \text{(Gain times effective Time scale)} \tag{2}$$

Because $g(x)$ decays rapidly, it is significantly non-zero only for small values of $x$ near $\tau$.

### 2 First-Order Approximation of the Convolution

The convolution of $f(t)$ and $g(t)$ is given by:

$$(f * g)(t) = \int_{-\infty}^{\infty} f(t-x)g(x)\,dx \tag{3}$$

Since $f(t)$ is a smooth, wide pulse, we can approximate $f(t-x)$ using a first-order Taylor series expansion around $t$:

$$f(t-x) \approx f(t) - xf'(t) \tag{4}$$

Substituting this back into the integral:

$$\begin{aligned}(f * g)(t) &\approx \int_{-\infty}^{\infty} [f(t) - xf'(t)]g(x)\,dx \\ &= f(t)\int_{-\infty}^{\infty} g(x)\,dx - f'(t)\int_{-\infty}^{\infty} xg(x)\,dx \\ &= G(f(t) - \tau f'(t))\end{aligned} \tag{5}$$

Recognizing that $f(t) - \tau f'(t)$ is the first-order Taylor expansion of $f(t-\tau)$, we conclude:

$$(f * g)(t) \approx G \cdot f(t-\tau) \tag{6}$$

Or put differently, the convolution of the slow pulse with the rapid kernel results in a function similar to the pulse in time $t - \tau$ multiplied by some number G.

### The Optimal Cross-Correlation of a Smooth Positive Function with the Product of the Function with Itself, Shifted in Time

#### 1 Introduction and the Gaussian Approximation

We are given a smooth positive function $f(t)$ with a single maximum. We want to find the time $T$ that maximizes the correlation between the product function $g(t) = f(t)f(t+\tau)$ and the shifted function $h(t) = f(t+T)$.

If $f(t)$ is symmetric, we can approximate it locally as a Gaussian:

$$f(t) \approx A \exp\left(-\frac{(t-\mu)^2}{2\sigma^2}\right) \tag{7}$$

The product $g(t) = f(t)f(t+\tau)$ becomes:

$$g(t) \approx A^2 \exp\left(-\frac{(t-\mu)^2 + (t+\tau-\mu)^2}{2\sigma^2}\right) \tag{8}$$

To find the peak of this product function, we set the derivative of the exponent to zero:

$$\frac{d}{dt}\left[(t-\mu)^2 + (t+\tau-\mu)^2\right] = 0$$
$$2(t-\mu) + 2(t+\tau-\mu) = 0$$
$$2t - 2\mu + \tau = 0 \implies t_{peak} = \mu - \frac{\tau}{2}$$

The shifted function $f(t+T)$ peaks when $t+T=\mu$, meaning $t_{peak} = \mu - T$. Aligning the peaks of both functions for maximum correlation gives:

$$\mu - \frac{\tau}{2} = \mu - T \implies T = \frac{\tau}{2} \tag{9}$$

For symmetric, bell-shaped functions, the optimal shift is exactly $T = \tau/2$.

#### 2 The General Case via Perturbation Expansion

To solve for a general, potentially asymmetric smooth function, we maximize the cross-correlation integral $C(T)$:

$$C(T) = \int_{-\infty}^{\infty} f(t)f(t+\tau)f(t+T)\,dt \tag{10}$$

Assuming $f(t)$ vanishes at $\pm\infty$, we set the derivative with respect to $T$ to zero:

$$\frac{dC}{dT} = \int_{-\infty}^{\infty} f(t)f(t+\tau)f'(t+T)\,dt = 0 \tag{11}$$

Assuming $\tau$ and $T$ are relatively small, we expand $f(t+\tau)$ and $f'(t+T)$ using Taylor series up to the second order:

$$f(t+\tau) \approx f(t) + \tau f'(t) + \frac{\tau^2}{2} f''(t)$$
$$f'(t+T) \approx f'(t) + T f''(t) + \frac{T^2}{2} f'''(t)$$

Substituting these into the integral yields:

$$\int_{-\infty}^{\infty} f\left(f + \tau f' + \frac{\tau^2}{2} f''\right)\left(f' + T f'' + \frac{T^2}{2} f'''\right) dt = 0 \tag{12}$$

#### 3 Integration by Parts

Expanding the product and dropping terms higher than second order leaves six terms to integrate (bounds are $-\infty$ to $\infty$):

$$\int \left[ f^2 f' + T f^2 f'' + \tau f f'^2 + \frac{T^2}{2} f^2 f''' + \frac{\tau^2}{2} f f' f'' + \tau T f f' f'' \right] dt = 0 \tag{13}$$

We define two shape parameters:

- **Sharpness:** $K = \int_{-\infty}^{\infty} f(t)[f'(t)]^2 \, dt$
- **Skewness:** $S = \int_{-\infty}^{\infty} [f'(t)]^3 \, dt$

Evaluating each term using integration by parts:

1. **Zeroth-order:** $\int f^2 f' \, dt = \left[\frac{1}{3} f^3\right]_{-\infty}^{\infty} = 0$
2. **$\tau$ term:** $\tau \int f f'^2 \, dt = K\tau$
3. **$T$ term:** $T \int f^2 f'' \, dt = -2KT$ (using $u = f^2, dv = f'' dt$)
4. **$\tau^2$ term:** $\frac{\tau^2}{2} \int f f' f'' \, dt = -\frac{1}{4} S\tau^2$ (using $u = f, dv = f' f'' dt$)
5. **$T^2$ term:** $\frac{T^2}{2} \int f^2 f''' \, dt = \frac{1}{2} ST^2$
6. **$\tau T$ term:** $\tau T \int f f' f'' \, dt = -\frac{1}{2} S\tau T$

Summing these terms yields the governing quadratic equation:

$$-2KT + \frac{1}{2} ST^2 + K\tau - \frac{1}{2} S\tau T - \frac{1}{4} S\tau^2 = 0 \tag{14}$$

For a symmetric function, $S = 0$, which yields $T = \frac{\tau}{2}$

#### 4 Solving the Quadratic Equation

We use perturbation theory to find a solution close to the symmetric case. Let $T = \frac{\tau}{2} + \delta$, where $\delta$ is a small correction. Substituting this into the quadratic equation:

$$\frac{1}{2} S \left(\frac{\tau}{2} + \delta\right)^2 - 2K \left(\frac{\tau}{2} + \delta\right) - \frac{1}{2} S\tau \left(\frac{\tau}{2} + \delta\right) + K\tau - \frac{1}{4} S\tau^2 = 0 \tag{15}$$

Expanding the terms:

$$\left(\frac{1}{8} S\tau^2 + \frac{1}{2} S\tau\delta + \frac{1}{2} S\delta^2\right) + (-K\tau - 2K\delta) + \left(-\frac{1}{4} S\tau^2 - \frac{1}{2} S\tau\delta\right) + K\tau - \frac{1}{4} S\tau^2 = 0 \tag{16}$$

The linear terms $(-K\tau + K\tau)$ and the mixed terms $(\frac{1}{2} S\tau\delta - \frac{1}{2} S\tau\delta)$ cancel out. Combining the $S\tau^2$ terms gives:

$$-2K\delta + \frac{1}{2} S\delta^2 - \frac{3}{8} S\tau^2 = 0 \tag{17}$$

Since $\delta$ is small (driven by $\tau^2$), the term $\delta^2$ is of fourth order ($\tau^4$) and can be dropped. We are left with:

$$-2K\delta - \frac{3}{8} S\tau^2 = 0 \implies \delta = -\frac{3S}{16K} \tau^2 \tag{18}$$

Substituting $\delta$ back gives the final general approximation:

$$T \approx \frac{\tau}{2} - \frac{3S}{16K} \tau^2 \tag{19}$$
